# Laboratory biomarkers associated with mortality in COVID-19 patients in Addis Ababa, Ethiopia

**DOI:** 10.1101/2023.05.20.23290268

**Authors:** Abay Sisay, Zerihun Woldesenbet, Anteneh Yalew, Aklilu Toma Shamenna, Asnake Worku, Abraham Tesfaye, Fentabil Getnet, Latera Tesfaye, Mohammed B. Hassen, Mulugeta Geleso, Veranyuy D Ngah, Perseverence Savieri, Alemnesh H. Mirkuze, Lovemore Sigwadhi, Adey Feleke Desta, Peter S Nyasulu

## Abstract

**Background:** Laboratory biomarkers are amongst the best imperative predictors of disease outcomes in hospital-admitted COVID-19 patients. Although data is available in this regard at a global level, there is a paucity of information in Ethiopia. Thus, this study aimed to assess the laboratory biomarkers association with death among COVID-19 patients in Ethiopia.

**Methods:** A health facility-based longitudinal study was conducted from 2020 to 2022 among RT-PCR-confirmed COVID-19 patients admitted and on treatment follow-up at COVID-19 treatment hospitals in Addis Ababa. A robust Poisson regression model was fitted to assess the association between demographic, clinical, and laboratory factors and death. Significance was determined at p<0.05, and variables with p□<□ 0.15 in bivariate analyses were included in the final multivariable models. Incidence rate ratio (IRR) with a 95% confidence interval (CI) was used to describe associations.

**Results:** Of the 2357 COVID-19 patients, 248 (10.5%) died. The median age of participants was 59 (IQR= 45-70) years, and the majority (64.9%) of them were male. Lower median RBC was observed among those who died at 4.58 (4.06-5.07) as compared to those who survived at 4.69 (4.23-5.12) whereas high median (IQR) WBC was a predictor of mortality with 11.2 (7.7-15.9). After adjusting for confounders, death was associated with age >74 years having adjusted incidence rate ratio [aIRR (95%CI): 2.46 (1.40-4.34)], and critical clinical situations [aIRR (95% CI): 4.04 (2.18-7.52)].

**Conclusion:** Our results demonstrate that abnormal liver function tests, abnormal white blood cells, age of the patients, and clinical status of the patients during admission are associated with unfavorable outcomes of COVID-19. Hence, timely monitoring of these laboratory results at the earliest phase of the disease was highly commendable.

## Introduction

The Coronavirus disease 2019 (COVID-19) is caused by the severe acute respiratory syndrome coronavirus 2 (SARS-CoV-2). As of January 2020, the World Health Organization (WHO) had declared it a Public Health Emergency of International Concern [1, 2]. On the African continent, more than 12.7 million cases with 258,122 deaths have been reported as of January 2023, far fewer than reported for other continents [1]. These low-reported numbers of infections and associated deaths in Africa were thought to reflect the relatively young population of Africa, climate conditions less favorable for disease transmission, reduced incidence of comorbidities, genetic factors coupled with immunological and socio-demographic aspects unique to Africa, and other anthropological factors [3-5]. Many studies have suggested that the relatively low observed rates could be a result of the poor documentation of the spread of SARS-CoV-2 in Africa [3, 6] and inadequate testing [7] and diagnostics [8].

The unprecedented spread of the virus across the globe and the abundance of genomic sequence data contributed by both research communities and public health laboratories have allowed for phylodynamic approaches to infer viral evolutionary rates, growth rates, and the estimation of the origin of specific outbreaks [9, 10]. SARS-CoV-2 has thus been the subject of numerous molecular epidemiological studies. Mostly, focusing on mutations in the viral spike protein, which is responsible for binding to the host angiotensin-converting enzyme 2 (ACE2) receptor and initiating the viral entry process. Such mutations confer viral phenotypes with increased fitness and pathogenicity [9, 11].

The viral variants that have been most intensively monitored by the WHO are those that have displayed phenotypes associated with increased infectivity, resistance to monoclonal antibody therapy, and evasion of the immune response. Upon emergence, viral lineages with these phenotypes often dominated the transmission and replacement of other co-circulating lineages [11, 12]. Accordingly, several variants of interest (VOIs) and variants of concern (VOCs) have been identified since December 2020. The Alpha/B.1.1.7 VOC was first identified in the United Kingdom in September 2020, the Beta VOC was first identified in South Africa in May 2020, the Gamma VOC was first identified in Brazil in November 2020, and the Delta VOC was first identified in India in late October 2020. The Omicron variant, however, was first identified in Botswana and South Africa in November 2021. The variant rapidly spread to the rest of the world and is currently the only variant still classified as a VOC [13, 14].

Laboratory biomarkers are biological evidence-based measurable indicators of some biological state or circumstance that can be summarized by undergoing various profiling experimental trials and tests to identify the unique biomarkers that correlate with the specific corresponding patients. These biomarkers could be very specific and can be used to monitor, assess the efficacy of treatments, and evaluate the prognosis of individual cases. Likewise, as part of pathophysiology and an indication of the dysfunction in distinctive organs and a crystal clear indication at the same time, due to their redundancy or non-specificity. It is very useful for monitoring COVID-19 cases and the correlation of their severity by timely risk stratification [5, 6].

Different countries, research institutes, universities, and other concerned organizations have been working to urgently design and recommend near-to-real-time laboratory biomarkers for monitoring the severity and treatment outcome of COVID-19 patients to augment the quality of life expectancy and their survival along with its alteration [7, 8].

Diverse laboratory biomarkers have been identified and drawn in as indicating substantial progress, level of severity, and disease outcome among COVID-19 patients: Cytopenia, misbalancing organ function tests, and electrolyte imbalance were among the top identified and more closely connected with unfavorable outcomes among hospitalized COVID-19 patients [5, 7].

Moreover biomarkers may enhance efforts through identification of cases which in turn may have helped reduce the burden of the COVID-19 in the early phase prior to the undesirable disaster and being used as an effective intervention for controlling the burden of the pandemic. However, there are extremely limited studies available in this regard in Ethiopia. Thus, this study aimed to describe the laboratory findings and their correlation with the prognosis in the outcome and severity of COVID□19 patients in Addis Ababa, Ethiopia.

## Materials and Methods

### Study design, period, and settings

A health facility-based cohort study was conducted among RT-PCR confirmed COVID-19 patients who were admitted and on treatment follow-up in a tertiary level hospital specifically designated for COVID-19 treatment centers in Addis Ababa, Ethiopia: Millennium Hall treatment center, Ekka Kotebe Hospital, and Yekatit 12 Hospital Medical College, encompassing a one-year cohort of COVID-19 patients’ clinical and laboratory findings from 2020 to 2022, a time between the first and second waves of the pandemic. Attached is the current epi plot, highlighting our sampling time (fig. 1).

**Fig 1.**
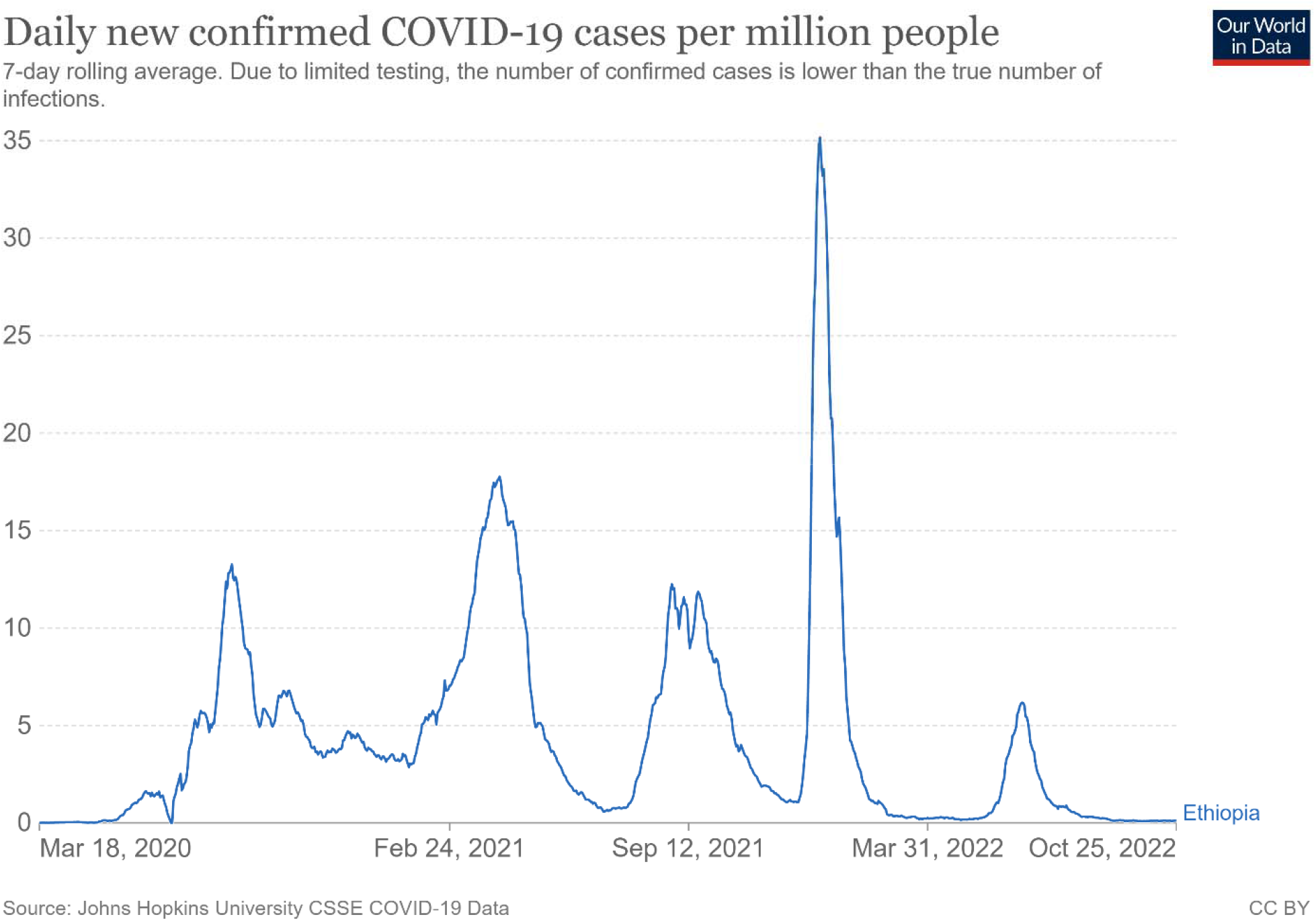
Epi plot of COVID-19 epidemic waves^@ Coronavirus (COVID-19) Cases - Our World in Data^

### Study Population and Sampling Method

We used prospective purposive continuous sampling and enrolled all eligible cases as they were admitted to the treatment center during the specified time frame. That means we did not use a specified sample size calculation. In this study, we excluded confirmed COVID-19-positive patients with some incomplete supportive data.

### Sample Collection and Laboratory testing

Venous blood was collected in serum separation gel tubes (SSGTs) for clinical chemistry test parameters and EDITA anti-coagulated tube for haematology analysis. We used very reputable analyzers: For Haematology - Beckman Coulter DxH 900 haematology analyzer. For Clinical Chemistry including electrolytes and hormones - Roche Cobas® 6000 automated analyzer series, manufactured in Germany. The laboratory tests: haematology, clinical chemistry, blood electrolyte, clotting factor, and organ function were done at Yekatit 12 Hospital Medical College, Millennium Hall, and Ekka Kotebe Hospital laboratories. These laboratories were among the laboratories accredited by the national laboratory for this purpose and get continuous quality improvement technical support.

### Quality Assurance

Appropriate training was given to the data collectors on how to get and assure valid data, as supported by active supervision to ensure the completeness and consistency of the data collection procedure. All clinical laboratory tests and interpretations were done following the analyzer manufacturers’ recommendations and approved documented standard operating procedures, S3. We always incorporated positive and negative quality controls into each laboratory test run. Moreover, data were double-entered and cross-checked and verified prior to analysis.

### Outcomes and predictor variables

Data collected included demographic data (age, sex), clinical characteristics (severity status), and clinical laboratory profiles: haematological (RBC, MCV, MCH, HCT, MPV, WBC, NE, LY, MO, EO, NRBC, etc), Clinical Chemistry, Organ function, hormonal function tests, and electrolyte analyte. The primary outcome of interest was the proportion of patients who die, irrespective of their severity status.

### Statistical Data Analysis and Interpretation

Continuous variables were expressed as medians with inter-quartile range (IQR) since all laboratory data were non-normally distributed. Categorical variables were expressed using frequencies and percentages. Chi-square and Wilcoxon-rank-sum tests were performed to test the population distribution associated with mortality among categorical variables and the medians for the continuous variable with p-values. A robust Poisson regression model was used to assess significant associations between demographic, clinical factors, laboratory variables, and death. Factors associated with death with a p-value < 0.15 in unadjusted univariate robust Poisson regression were included in a multivariable model, to identify predictor variables associated with death. Due to the high prevalence of mortality, the logistic regression was overestimating the effect measure with large standard errors, resulting in wide confidence intervals. Therefore, a robust Poisson regression model was used. Adjusted incidence rate ratios and their 95% CIs were used as a measure of association. Factors associated with mortality with a p-value < 0.05 were considered statistically significant. All statistical analyses were performed using Stata (V.17, Stata Corp, College Station, Texas, USA) and R (V, 4.2.1, R Core Team) with R Studio (V. 2022.07.0, R Studio Team) statistical software.

### Operational Definition [9]

#### Mild COVID-19

characterized by fever, malaise, cough, upper respiratory symptoms, and/or less common features of COVID-19 (headache, loss of taste or smell, etc.)

#### Moderate COVID-19

patients with lower respiratory symptom/s. They may have infiltrates on chest X-ray. These patients can maintain oxygenation on room air.

#### Severe COVID-19

patients with oxygen saturation < 90% on room air; ° in adults, signs of severe respiratory distress (accessory muscle use, inability to complete full sentences, respiratory rate > 30 breaths per minute), and, in children, very severe chest wall in drawing, grunting, central cyanosis, or presence of any other general danger signs (inability to breastfeed or drink, lethargy or reduced level of consciousness, convulsions) in addition to the signs of pneumonia.

#### Test abnormalities

were defined as an elevation or reduction of test results against the specified manufacturer’s recommendation.

#### Critical COVID-19

patients suffering from acute respiratory distress syndrome (ARDS), sepsis, septic shock, or other illnesses requiring life-sustaining therapies such as mechanical ventilation (invasive or non-invasive) or vasopressor therapy.

## Results

Of the 2357 patients with COVID-19 studied, 248 (10.5%) died, 1652 (70.2%) had severe COVID-19 disease at admission, and 1613 (68.4) were male and more severely affected than females. The majority of the cases had abnormal hematology findings, liver function tests, and electrolyte findings. The median RBC and WBC were 4.67 (4.205- 5.11) and 9.2 (6.3 - 13.6), respectively. Lower median RBC was observed among those who died at 4.58 (4.06-5.07) as compared to those who survived at 4.69 (4.23-5.12) whereas high median (IQR) WBC was a predictor of mortality with 11.2 (7.7-15.9). Alike, higher median (IQR) BILL and UREA were a predictor of mortality with 0.2105 (.133-.347) and 34.7 (25.5-54.2), respectively (Table 1).

**Table 1.** Demographic, clinical, and laboratory characteristics associated with mortality among COVID-19 patients in Addis Ababa

| Characteristics | Sample Size | Total (n = 2357) | Survived (n<sub>1</sub> = 2109) | Died (n<sub>2</sub> = 248) |  |
| --- | --- | --- | --- | --- | --- |
|  |  | Median (IQR) or n (%) | Median (IQR) or n (%) | Median (IQR) or n (%) | P-value |
| Age in years | 1671 | 59 (45- 70) | 58 (45-70) | 65 (50-74) | <0.001 |
| Sex | 2357 |  |  |  | 0.208 |
| Female |  | 744 (31.6) | 657 (31.2) | 87 (35.1) |  |
| Male |  | 1613 (68.4) | 1452 (68.8) | 161 (64.9) |  |
| Severity status | 2357 |  |  |  | <0.001 |
| Mild |  | 152 (6.4) | 138 (6.6) | 14 (5.6) |  |
| Moderate |  | 442 (18.8) | 420 (19.9) | 22 (8.9) |  |
| Severe |  | 1652 (70.1) | 1488 (70.7) | 164 (66.1) |  |
| Critical |  | 111 (4.7) | 60 (2.8) | 48 (19.4) |  |
| **Laboratory parameters (Hematology and analytes)** |  |  |  |  |  |
| RBC | 2040 | 4.67 (4.205- 5.11) | 4.69 (4.23-5.12) | 4.58 (4.06-5.07) | 0.041 |
| MCV | 2040 | 89.1 (86.1- 92.4) | 89 (86.1-92.3) | 89.8 (86.3-93.1) | 0.065 |
| MCH | 2040 | 33.6 (32.9- 34.3) | 33.6 (32.9-34.3) | 33.4 (32.7-34.1) | 0.013 |
| HCT | 2040 | 41.7 (37.8 – 45.2) | 41.8 (37.9-45.3) | 41 (37-44.8) | 0.087 |
| MPV | 2040 | 8.6 (7.9 – 9.4) | 8.6 (7.9-9.3) | 8.5 (7.9-9.6) | 0.920 |
| WBC | 2039 | 9.2 (6.3 – 13.6) | 8.9 (6.2-13.15) | 11.2 (7.7-15.9) | <0.001 |
| Platelet | 2038 | 240 (176-322) | 240 (176-321) | 246 (169-340) | 0.653 |
| NE | 2019 | 7.9 (4.7- 12.4) | 7.6 (4.6-12.1) | 10.1 (6.4-14.8) | <0.001 |
| LY | 2019 | 0.6 (0.4- 1.2) | 0.6 (0.4-1.2) | 0.5 (0.3-0.8) | <0.001 |
| MO | 2019 | 0.4 (0.3- 0.6) | .4 (.3-.6) | .4 (.3-.6) | 0.420 |
| EO | 2 016 | 0 (0- 0.1) | 0 (0-0.1) | 0 (0-0.1) | 0.120 |
| NRBC | 1 999 | 0.1 (0.02- 0.2) | 0.1 (.02-.2) | 0.1 (.07-.2) | 0.240 |
| CRE | 1 926 | 0.78 (0.63- 0.98) | 0.78 (0.63-0.98) | 0.79 (0.62-1.01) | 0.720 |
| UREA | 1 868 | 30.9 (22.45- 46.4) | 30.3 (22.1-45) | 34.7 (25.5-54.2) | <0.001 |
| ALP | 1465 | 83.6 (65-114.6) | 83.05 (65-114.6) | 87 (65-114.6) | 0.191 |
| HGB | 1 861 | 14 (12.5- 15.3) | 14.1 (12.6-15.3) | 13.85 (12.05-15) | 0.100 |
| ALT | 1850 | 32 (20.5-57.1) | 32.1 (20.65-56.65) | 31.1 (19.7-62.0) | 0.695 |
| AST | 1847 | 32.2 (22.3-49.7) | 32.1 (22.2-49.5) | 32.8 (24.1-49.9) | 0.450 |
| Na | 1793 | 137 (134-140) | 137 (134-140) | 137 (133-140) | 0.289 |
| CL | 1 764 | 100 (96.6- 103.6) | 100.1 (96.7-103.6) | 99.7 (95.7-103.6) | 0.640 |
| K | 1 738 | 4.285 (3.91- 4.7) | 4.27 (3.9-4.68) | 4.4 (3.99-4.79) | 0.025 |
| MG | 867 | 0.85 (0.76- 0.96) | 0.85 (0.76-0.96) | 0.865 (0.77-0.96) | 0.750 |
| CA | 815 | 2.03 (1.91- 2.15) | 2.03 (1.91-2.15) | 2.02 (1.9-2.14) | 0.920 |
| DBIL | 752 | 0.186 (0.119- 0.289) | 0.183 (0.118-0.282) | 0.2105 (.133-.347) | 0.046 |
| TBILI | 693 | 0.416 (0.288- 0.604) | 0.416 (0.286-0.599) | 0.422 (0.305-0.619) | 0.380 |
| APTT | 510 | 27.7 (24.7- 31.4) | 27.7 (24.7-31.3) | 28.7 (25.2-32.4) | 0.170 |
| INR | 489 | 1.14 (1.01 - 1.31) | 1.14 (1.01- 1.31) | 1.135 (1.02- 1.26) | 0.805 |
| CHOL | 326 | 130.1 (102.4- 164.1) | 132.8 (104.1- 166.1) | 114.5 (93.1- 150.9) | 0.033 |
| ALB | 291 | 1.89 (1.53- 2.37) | 1.91 (1.55- 2.41) | 1.73 (1.5- 2.22) | 0.186 |
| HDL | 236 | 31.65 (24.45- 39.65) | 31.2 (24.5- 39.7) | 34 (24.3- 39.6) | 0.458 |
| TG | 229 | 141.6 (108- 206) | 144 (108- 223) | 128 (100- 160) | 0.028 |
| LDL | 167 | 77.4 (47.6- 103.2) | 80.25 (51.1- 104.3) | 61.3 (42- 91.8) | 0.293 |
| CKMB | 44 | 2.04 (1.4- 3.2) | 2.08 (1.43- 3.4) | 1.37 (1.23- 2.4) | 0.999 |
| GLU | 39 | 135 (96- 218.4) | 125.05 (94.2- 189.8) | 212.9 (136.2- 261.3) | 0.081 |
| TSH | 31 | 0.77 (0.28- 1.69) | 0.78 (0.245- 2.04) | 0.582 (0.39- 1.6) | 0.953 |
| T4 | 31 | 98.21 (11.92- 111.2) | 98.79 (13.61- 115.985) | 93.51 (11- 99.21) | 0.953 |
| T3 | 25 | 1.76 (1.2- 2.12) | 1.78 (1.2- 2.12) | 1.71 (0.706- 2.47) | 0.941 |
| TP | 10 | 2.17 (0.67- 5.65) | 2.21 (0.67- 5.65) | 2.02 (2.02- 2.02) | 0.999 |

*\*All these lab characteristics were non-normally distributed using the Shapiro–Francia test of normality. As a result, we presented a median with an interquartile range [median (lower quartile – upper quartile)]. Abbreviations: RBC: red blood cells; etc.*

### Factors Associated with the unaffordable outcome of COVID-19 in Addis Ababa, Ethiopia

The COVID-19 pandemic constantly increased, overwhelming the treatment sites and the attending health professionals in assisting the admitted COVID-19 patients, as they judiciously wanted to have a successful treatment recovery rate. To realize this, we identified and documented the most contributing factors associated with the clinical outcomes of hospitalized COVID-19 patients. After adjusting for confounders, we identified associated predictors with poor outcomes among studied COVID-19 patients. Higher death among COVID-19 patients of older age (Age >74 yrs.: aIRR 2.46; 95% CI: 1.40-4.34, P =0.002) were revealed than the younger COVID-19 patients. Likewise, patients who were in critical clinical situations at the time of admission were four times more likely to have unfavorable COVID-19 outcomes (95% CI: 2.18-7.52, p<0.001). The detail is depicted in Table 2, Fig. 2, and S3.

**Table 2.**
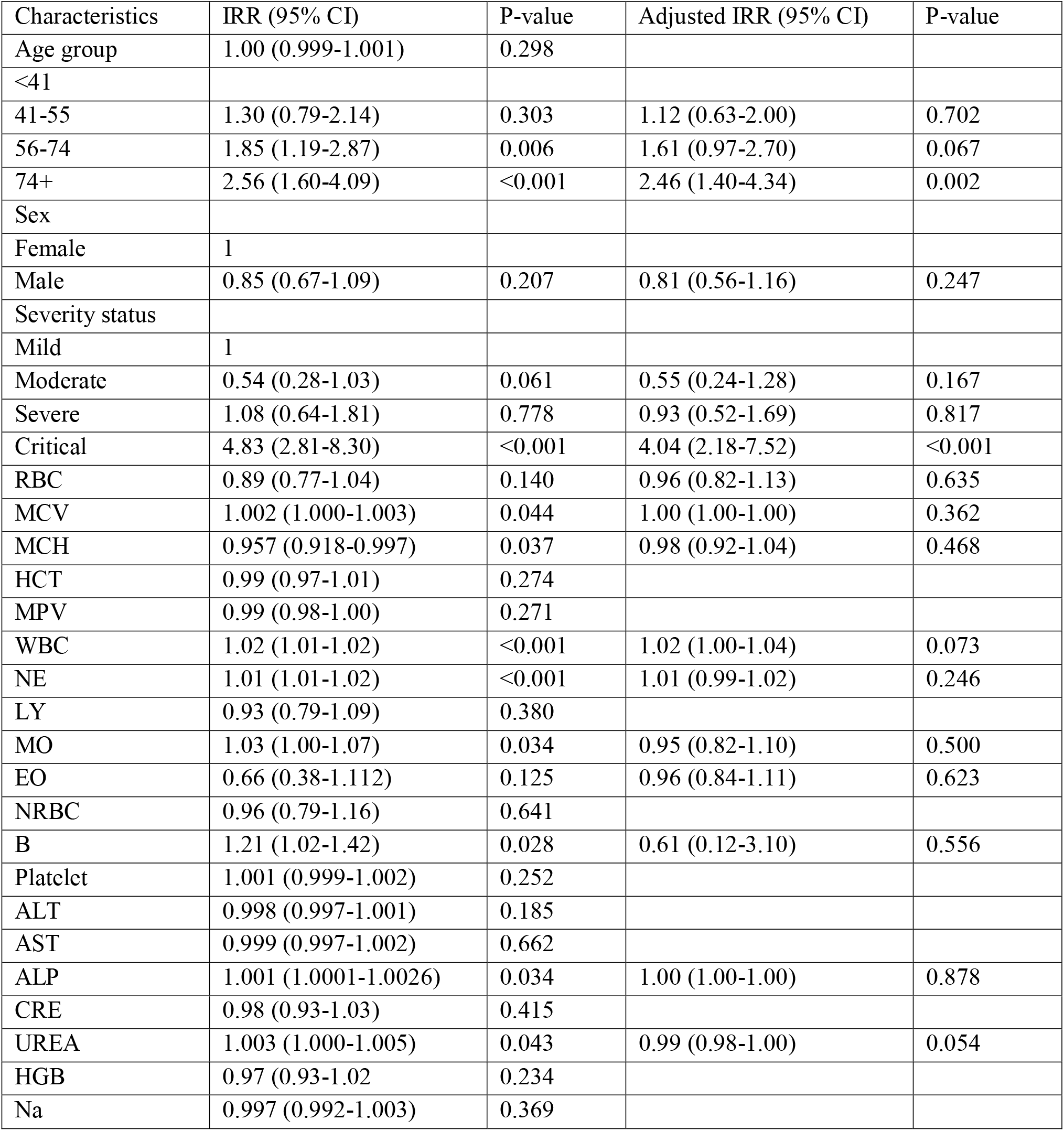
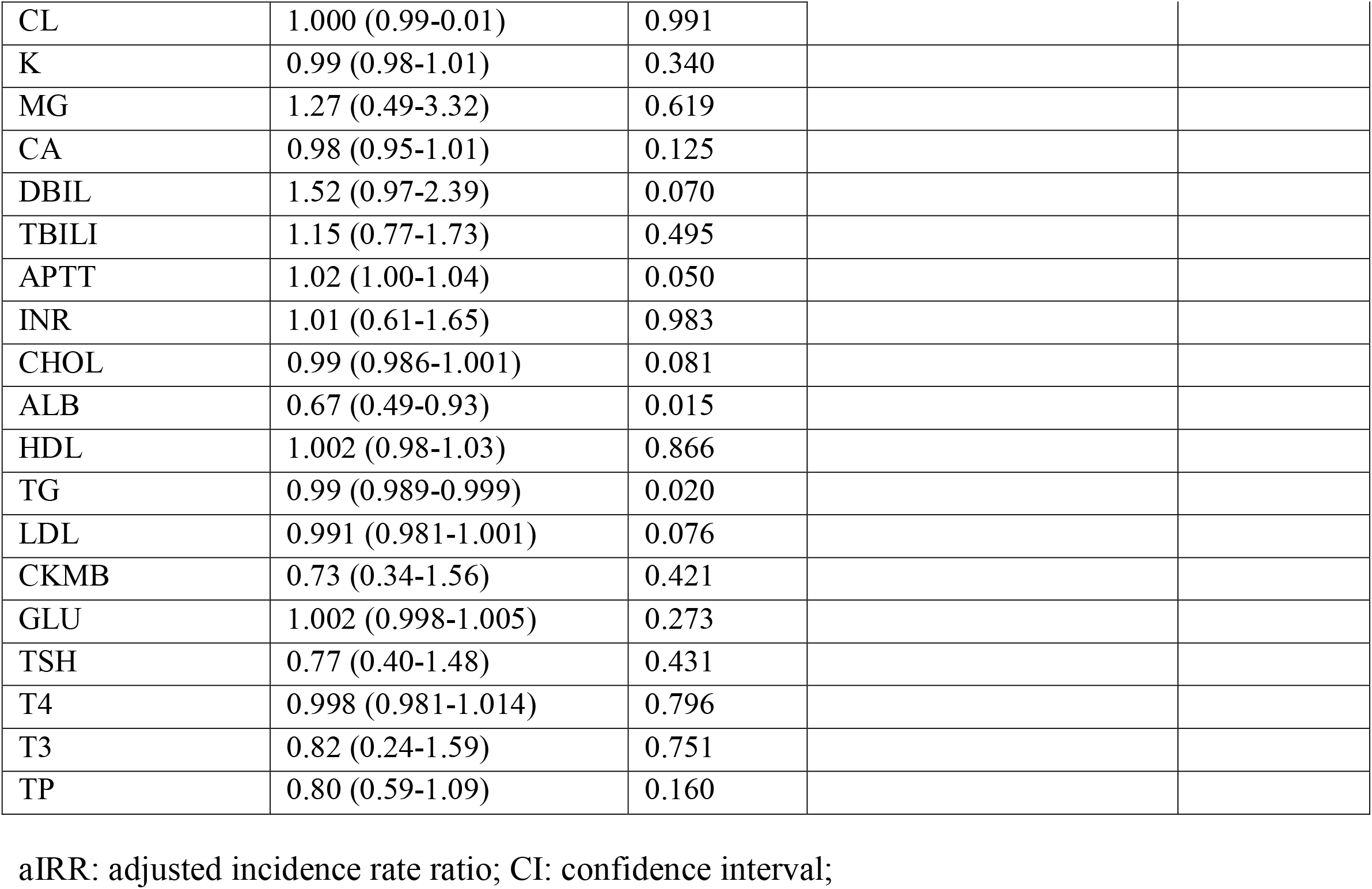
Robust Poisson Regression analysis among laboratory characteristics associated with mortality among COVID-19 patients in Addis Ababa.

| Characteristics | IRR (95% CI) | P-value | Adjusted IRR (95% CI) | P-value |
| --- | --- | --- | --- | --- |
| Age group | 1.00 (0.999-1.001) | 0.298 |  |  |
| <41 |  |  |  |  |
| 41-55 | 1.30 (0.79-2.14) | 0.303 | 1.12 (0.63-2.00) | 0.702 |
| 56-74 | 1.85 (1.19-2.87) | 0.006 | 1.61 (0.97-2.70) | 0.067 |
| 74+ | 2.56 (1.60-4.09) | <0.001 | 2.46 (1.40-4.34) | 0.002 |
| Sex |  |  |  |  |
| Female | 1 |  |  |  |
| Male | 0.85 (0.67-1.09) | 0.207 | 0.81 (0.56-1.16) | 0.247 |
| Severity status |  |  |  |  |
| Mild | 1 |  |  |  |
| Moderate | 0.54 (0.28-1.03) | 0.061 | 0.55 (0.24-1.28) | 0.167 |
| Severe | 1.08 (0.64-1.81) | 0.778 | 0.93 (0.52-1.69) | 0.817 |
| Critical | 4.83 (2.81-8.30) | <0.001 | 4.04 (2.18-7.52) | <0.001 |
| RBC | 0.89 (0.77-1.04) | 0.140 | 0.96 (0.82-1.13) | 0.635 |
| MCV | 1.002 (1.000-1.003) | 0.044 | 1.00 (1.00-1.00) | 0.362 |
| MCH | 0.957 (0.918-0.997) | 0.037 | 0.98 (0.92-1.04) | 0.468 |
| HCT | 0.99 (0.97-1.01) | 0.274 |  |  |
| MPV | 0.99 (0.98-1.00) | 0.271 |  |  |
| WBC | 1.02 (1.01-1.02) | <0.001 | 1.02 (1.00-1.04) | 0.073 |
| NE | 1.01 (1.01-1.02) | <0.001 | 1.01 (0.99-1.02) | 0.246 |
| LY | 0.93 (0.79-1.09) | 0.380 |  |  |
| MO | 1.03 (1.00-1.07) | 0.034 | 0.95 (0.82-1.10) | 0.500 |
| EO | 0.66 (0.38-1.112) | 0.125 | 0.96 (0.84-1.11) | 0.623 |
| NRBC | 0.96 (0.79-1.16) | 0.641 |  |  |
| B | 1.21 (1.02-1.42) | 0.028 | 0.61 (0.12-3.10) | 0.556 |
| Platelet | 1.001 (0.999-1.002) | 0.252 |  |  |
| ALT | 0.998 (0.997-1.001) | 0.185 |  |  |
| AST | 0.999 (0.997-1.002) | 0.662 |  |  |
| ALP | 1.001 (1.0001-1.0026) | 0.034 | 1.00 (1.00-1.00) | 0.878 |
| CRE | 0.98 (0.93-1.03) | 0.415 |  |  |
| UREA | 1.003 (1.000-1.005) | 0.043 | 0.99 (0.98-1.00) | 0.054 |
| HGB | 0.97 (0.93-1.02) | 0.234 |  |  |
| Na | 0.997 (0.992-1.003) | 0.369 |  |  |
| CL | 1.000 (0.99-0.01) | 0.991 |  |  |
| K | 0.99 (0.98-1.01) | 0.340 |  |  |
| MG | 1.27 (0.49-3.32) | 0.619 |  |  |
| CA | 0.98 (0.95-1.01) | 0.125 |  |  |
| DBIL | 1.52 (0.97-2.39) | 0.070 |  |  |
| TBILI | 1.15 (0.77-1.73) | 0.495 |  |  |
| APTT | 1.02 (1.00-1.04) | 0.050 |  |  |
| INR | 1.01 (0.61-1.65) | 0.983 |  |  |
| CHOL | 0.99 (0.986-1.001) | 0.081 |  |  |
| ALB | 0.67 (0.49-0.93) | 0.015 |  |  |
| HDL | 1.002 (0.98-1.03) | 0.866 |  |  |
| TG | 0.99 (0.989-0.999) | 0.020 |  |  |
| LDL | 0.991 (0.981-1.001) | 0.076 |  |  |
| CKMB | 0.73 (0.34-1.56) | 0.421 |  |  |
| GLU | 1.002 (0.998-1.005) | 0.273 |  |  |
| TSH | 0.77 (0.40-1.48) | 0.431 |  |  |
| T4 | 0.998 (0.981-1.014) | 0.796 |  |  |
| T3 | 0.82 (0.24-1.59) | 0.751 |  |  |
| TP | 0.80 (0.59-1.09) | 0.160 |  |  |

aIRR: adjusted incidence rate ratio; CI: confidence interval;

**Figure 2a:**
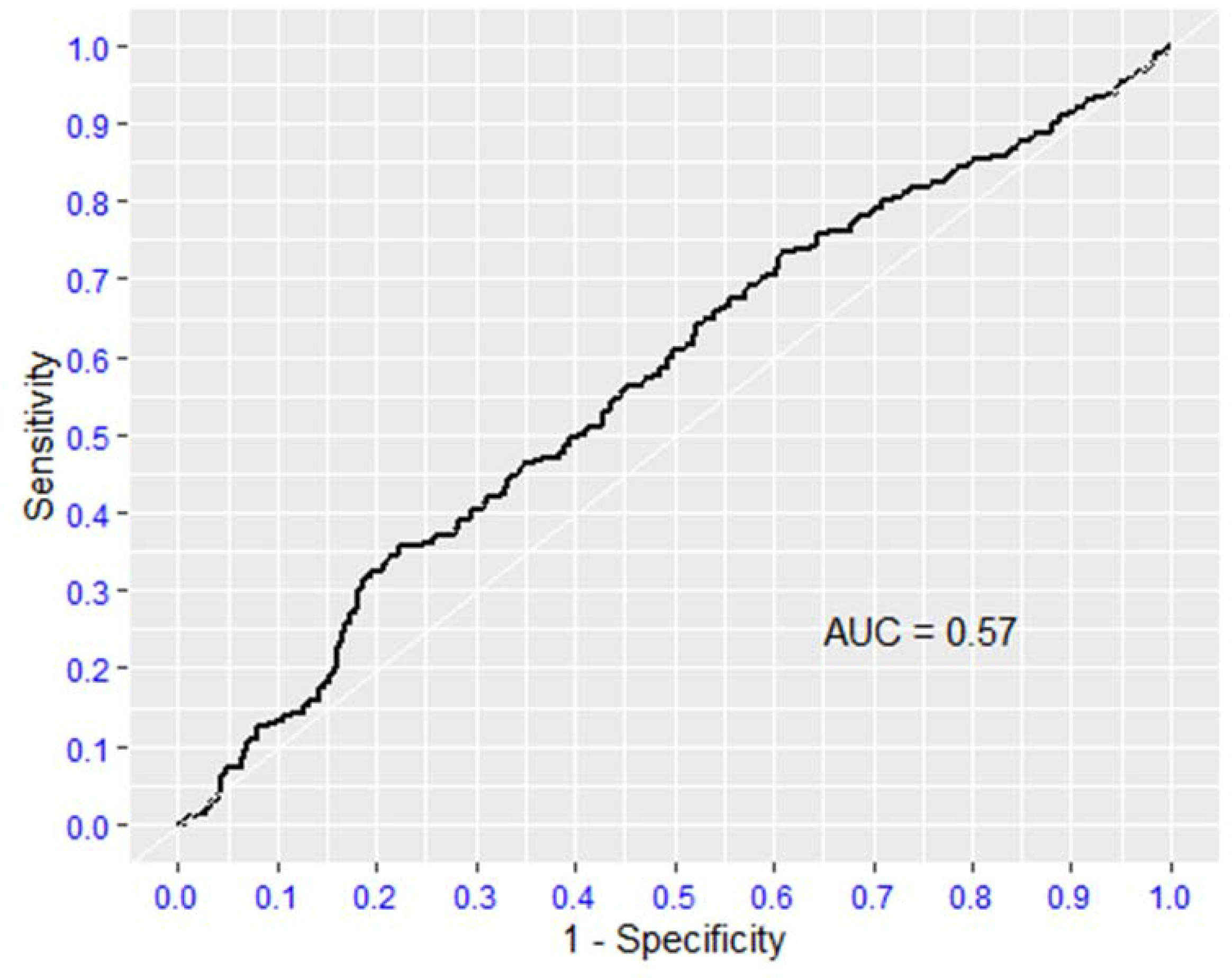
Roc curve Urea

**Figure 2b:**
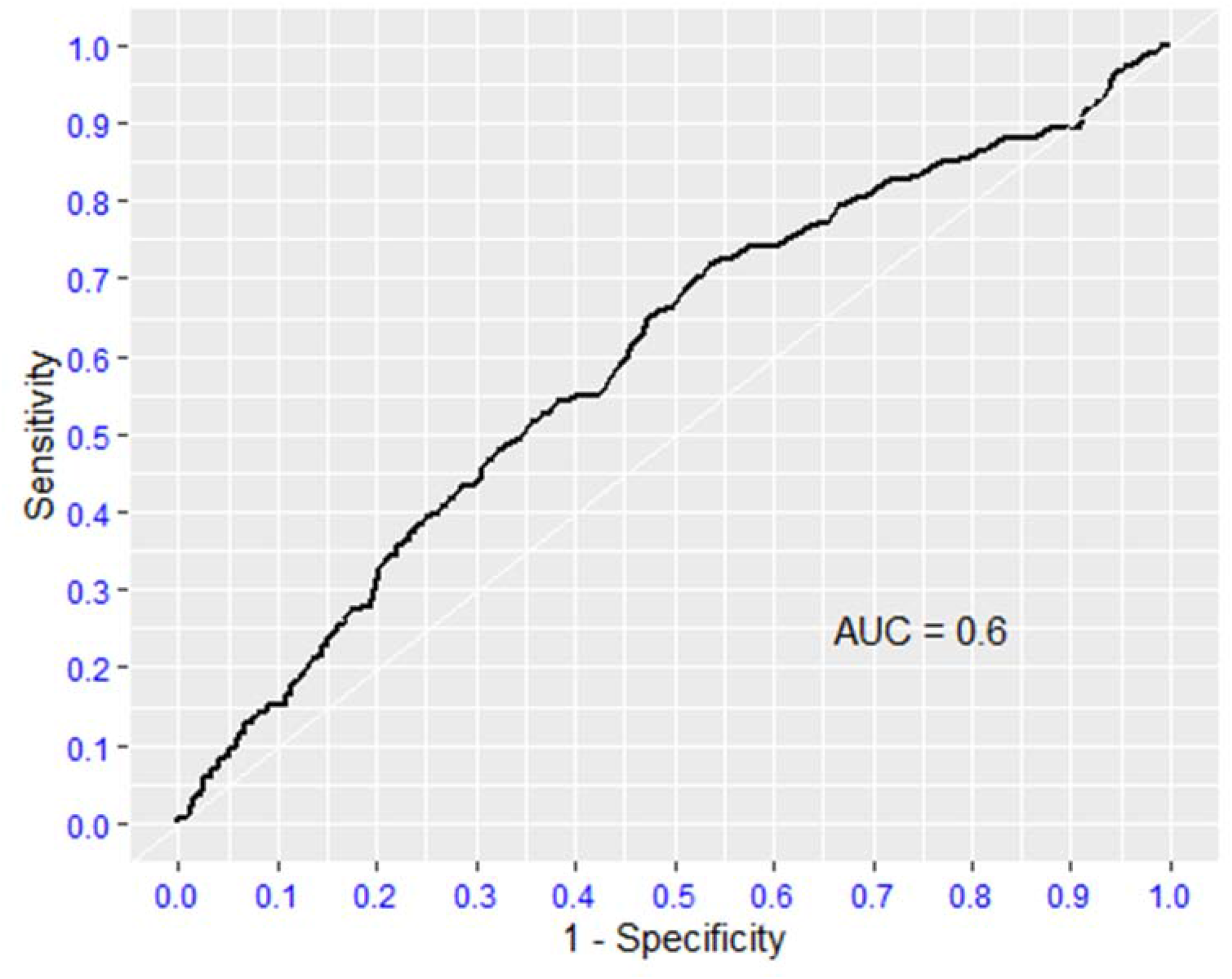
Roc curve WBC

## Discussion

Investigating and understanding the laboratory biomarkers in the human health medical process favors the early initiation of the most appropriate stratification therapy and increases the chances of survival. Although many studies and pieces of literature have reported as many laboratory biomarkers as predictors of severity related to COVID-19 severity globally, there is limited evidence in Ethiopia. Hence, we investigated the clinical laboratory profiles of COVID-19 RT-PCR-confirmed hospitalized patients in Addis Ababa, Ethiopia.

Thus, after adjusting for other covariates in our study, being in the critical status of COVID-19 was found to be associated with 4.104 times increased risk and ended up with unfavorable outcomes, as clearly recognized in different studies conducted in China, the United States, and Europe **[10-12]**. There’s a high possibility that the hyper-triggering and greater destabilization of metabolic reactions and physiologic effects might lead to an imbalance of laboratory biomarkers, which is a precarious physiological response in patients with severe COVID-19.

The probability of a destitute and deprived treatment outcome increases with the age of the patients during enrollment. Older patients, especially those 60 years and older, were more likely than younger patients, to have poor treatment results according to studies from China **[13]** and Saudi Arabia **[14]**. This might be associated with the body’s immunity status weakening with age. Likewise, older COVID-19 patients were more susceptible to severe disease and pitiable treatment outcomes from COVID-19. Patients having and presenting one or more certain underlying clinical situations are subject to acquiring the disease and being at higher risk. Additionally, being unvaccinated or not being up to date on COVID-19 vaccinations also increases the risk of severe COVID-19 outcomes. This could be more related to the fact that they were prone to many underlined diseases and that mired their medical status **[15]**.

The present result shows that adult males were more severely affected than females. Even though this might be subjected to a further large-scale population-based study, it could be illuminated by the dissimilarity of smoking rates, that more smoking can expose continuous accumulation of white blood cells and excretion of many pro-inflammatory cytokines against the provoking sense of the body’s immunity. Moreover, this could be more related to ACE2 and Oestrogen concentration levels. Accordingly, we have given due attention to those clients **[16, 17]**.

In light of our recent study, more than 10.5% of COVID-19 patients were dead among those enrolled in treatment centers and had poor treatment outcomes, which is concordant and comparable to Wuhan (11.73%) **[18]** and much lower than a report from Belgium (29.9%) **[19**] and China (28.27%) **[20]**. These incongruences could be the result of differences in study time, which are mostly related to the circulating variant of concerns’ (VOC) virulence, pathogenicity, and treatment care quality. Furthermore, this could be related to differences in treatment facilities and settings.

Disproportionately elevated levels of urea among the COVID-19 cases an indicator of acute renal injury were 0.99 (0.98-1.00, p=0.054) less likely to develop unfavorable disease outcomes.

An unfavorable and imperative prognosticator of respiratory failure among COVID-19 patients has been linked to more severe disease.

## Conclusion and Recommendation

Laboratory biomarkers were much more connected with disease severity and their outcome among COVID-19-positive patients. Given this, we have enrolled and examined more than 2000 COVID-19-positive clients and noted that timely laboratory biomarkers investigation and management can reverse the unfavorable disease outcome. The current findings might be a good indication and foundation data for expediting preparations for the upcoming surge of COVID-19 and related diseases caused by emerging pathogens.

## Data Availability

All data sets used in this study are included in this manuscript.

## Authors’ contributions

Conceptualization: AS, ZW, ATe(Abraham Tesfaye), AFD

Data curation: AS, AY, AT, ATe, LT, MB, MG, VDN, LS, PN, AFD

Formal analysis: AY, AT, ATe, LT, MB, MG, VDN, PS, LS, PN, AFD

Investigation: AS, ZW, AY, AT, LT, MB, MG, VDN, LS, PN, AFD

Methodology: AS, AY, AT, ATe, LT, MB, MG, VDN, PS, LS, PN, AFD

Project administration: AS, ZW, AFD

Resources: AS, ZW, AFD

Software: AY, AT, LT, MB, MG, VDN, LS, PN, AFD

Supervision: AS, ZW, AW, ATe, MB, LS, PN, AFD

Validation: AS, AY, AT, ATe, FG, LT, MB, MG, VDN, PS, LS, PN, AFD

Visualization: AS, AY, AT, AW, ATe, FG, LT, MB, MG, VDN, LS, PN, AFD

Writing original draft: AS, ZW, AY, AT, AW, ATe, FG, LT, MB, MG, VDN, PS, AHM, LS, PN, AFD

Writing review & editing: AS, ZW, AY, AT, AW, FG, LT, MG, VDN, PS, AHM, LS, PN, AFD

## Conflicts of interest

We, the authors declare that we have no known competing financial interests or personal relationships that could have appeared to influence the work reported in this paper.

## Data Availability Statement

All data sets used in this study are included in this manuscript.

## Funding

This work was supported and financed by Yekatit 12 Hospital Medical College, with project title, evaluation and determining laboratory profile and biomarkers among COVID-19 patients in Addis Ababa, Ethiopia, Ref # 75/20 and Addis Ababa University through an adaptive problem-solving project, with project title, Evaluation and Validation of the Diagnostic Performance of SARS-CoV-2 Rapid Test for the detection of Novel Corona Virus, Ref #-PR/5.15/590/12/20. No additional external funding was received. The funders are not involved in study design, data collection, and analysis, the decision to publish, or the preparation of the manuscript. The contents are virtuously the responsibility of the authors and do not represent or reflect the funder’s view.

## Ethical consideration

The research protocol had been getting approval from different institutional ethics and research committees prior to the analysis. Accordingly, it was approved by the IRB of the College of Health Sciences, Addis Ababa University IRB committee (IRB # 029/20/Lab), IRB of the Department of Medical Laboratory Sciences, Addis Ababa University (reference #-MLS/174/21), IRB office of Addis Ababa City Administration Health Bureau, AAPHREML (Reference #-AAHB/4039/227) and also by Yekatit 12 Hospital Medical College (IRB protocol # 75/20) and also reviewed and approved by the Ekka Kotebe hospital IRB protocol # Eka-150-5-4). Study participants consented/assented in writing to sample storage and subsequent use of their samples, and a waiver of consent was provided by the institutional review board for the use of leftover samples. Moreover, this study was conducted per the Helsinki Declaration’s general guiding principles and regulations.

## Acknowledgments

We are very grateful and acknowledge Addis Ababa University, Yekatit 12 Hospital Medical College, Ekka Kotebe Hospital, Millennium Hall COVID-19 Treatment Center, and Addis Ababa Health Bureau for their grant and their strong support and assistance in accessing diverse resources used in the study, including their institutional ethical approval.

## Annex, Supplementary Files

**S1.**
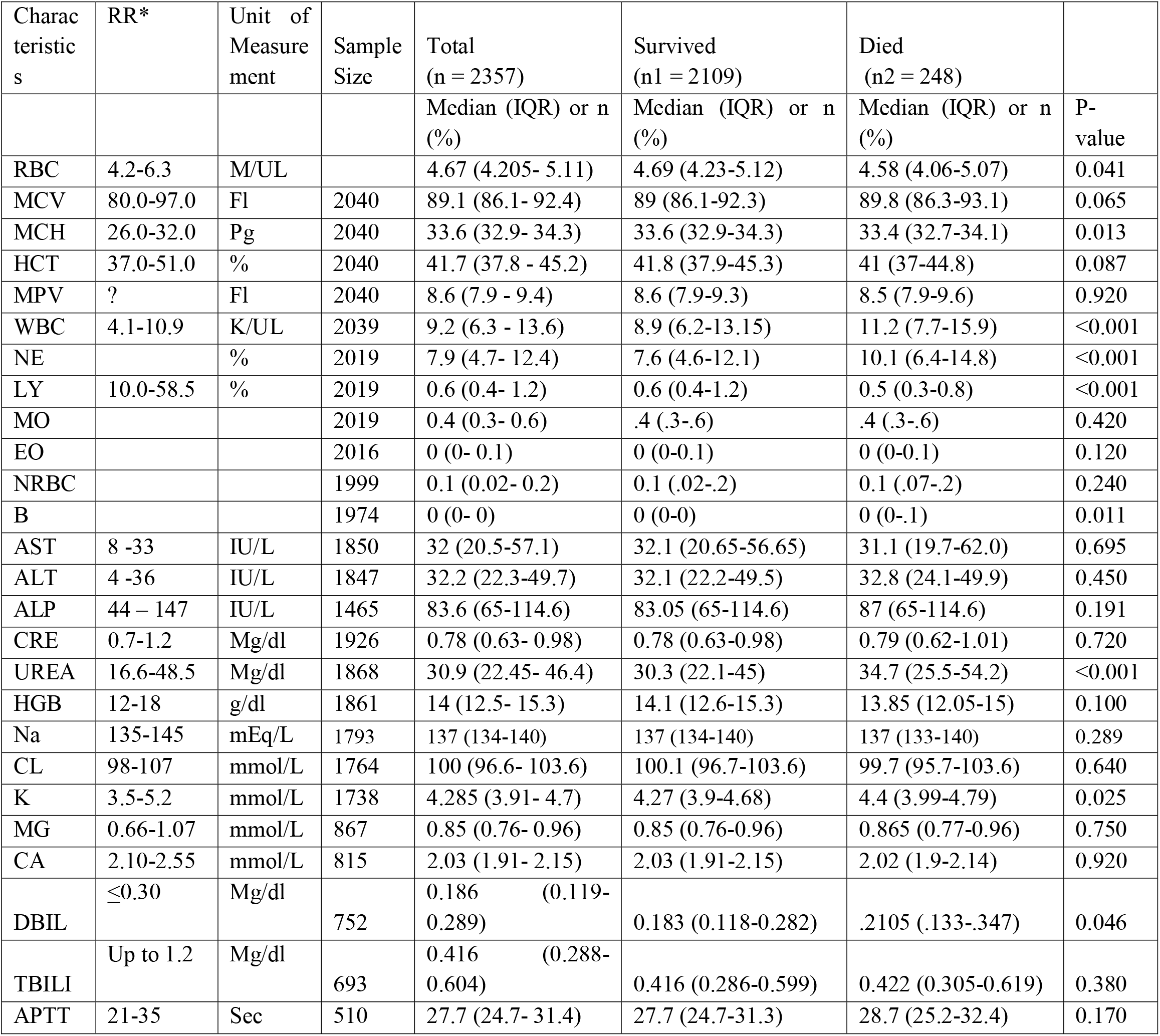
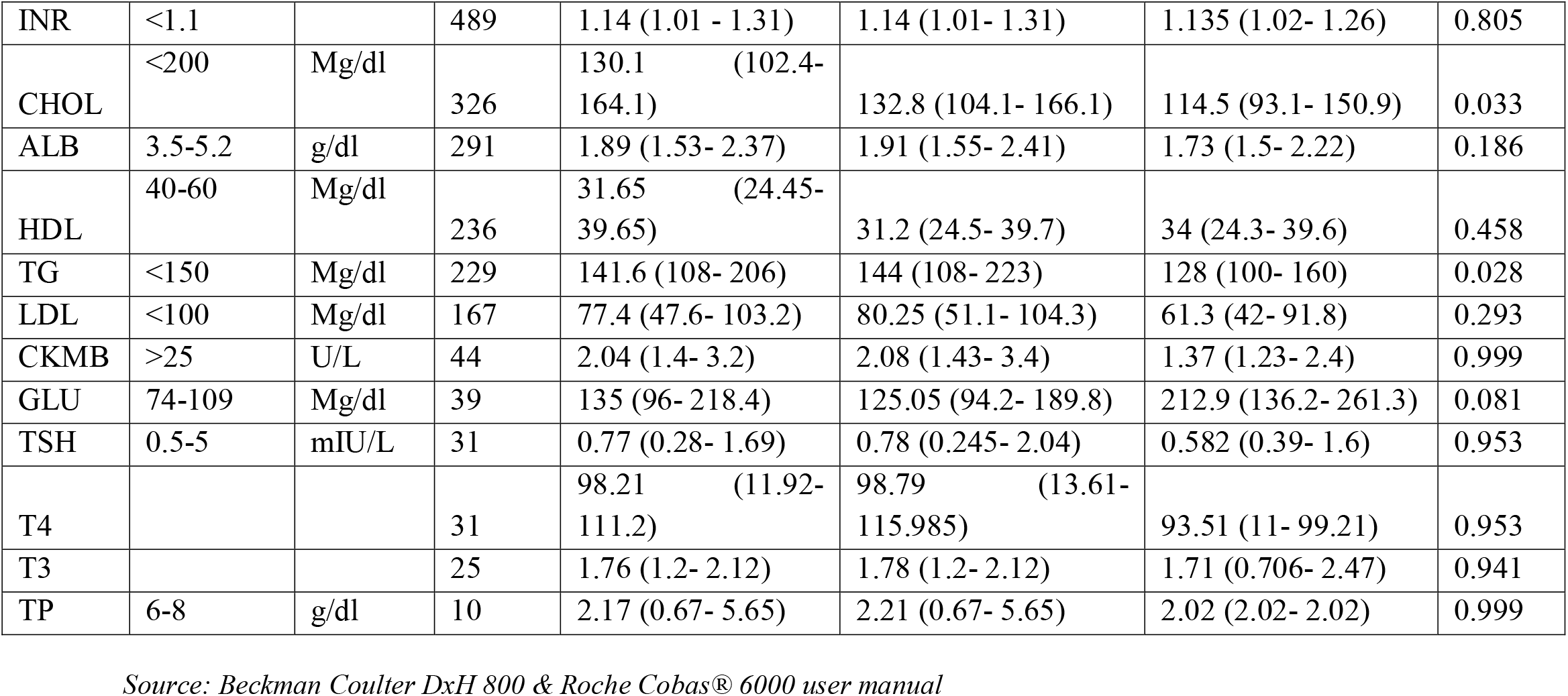
Our Laboratory findings VS reference range of the variables.

**S2: Supplementary file 2.**
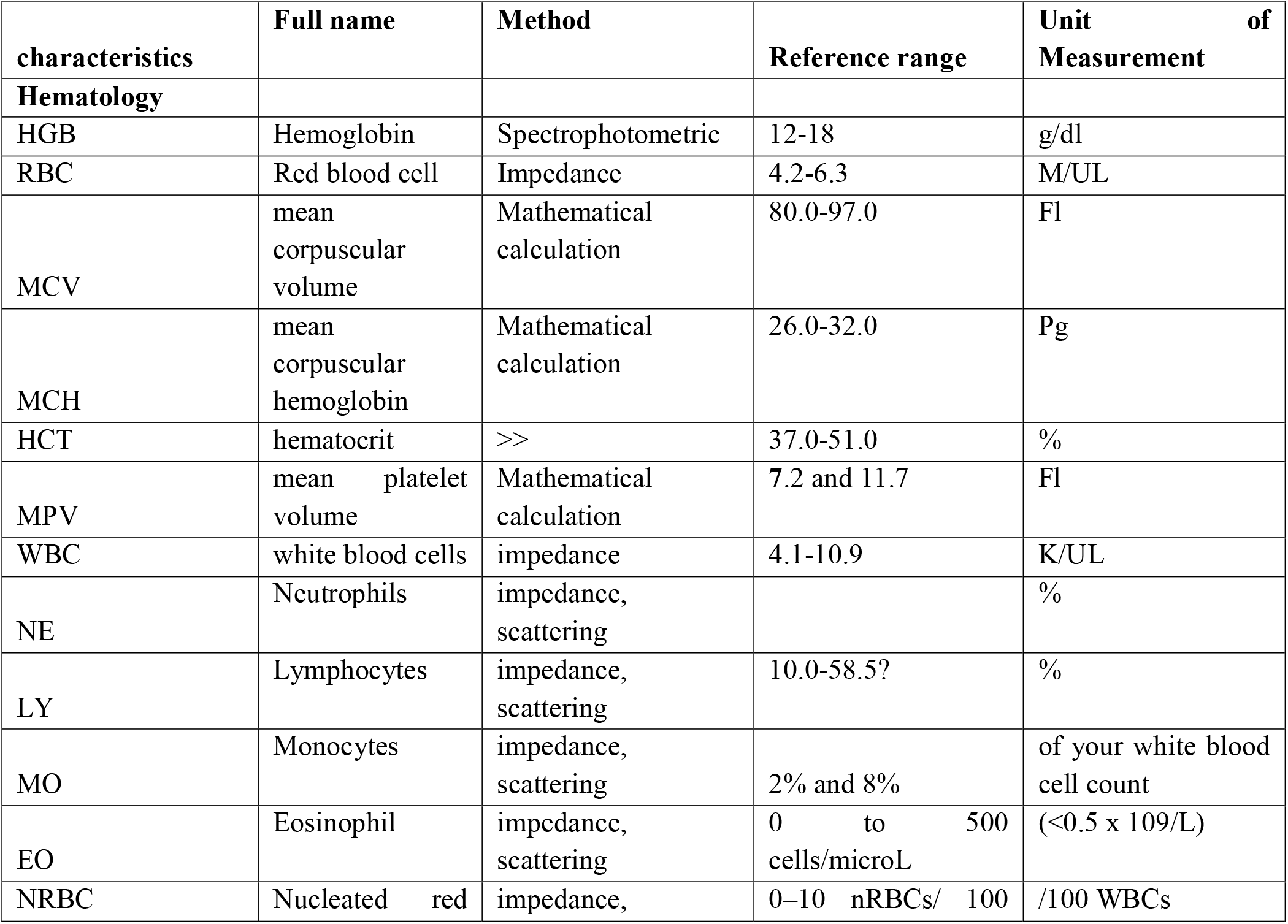
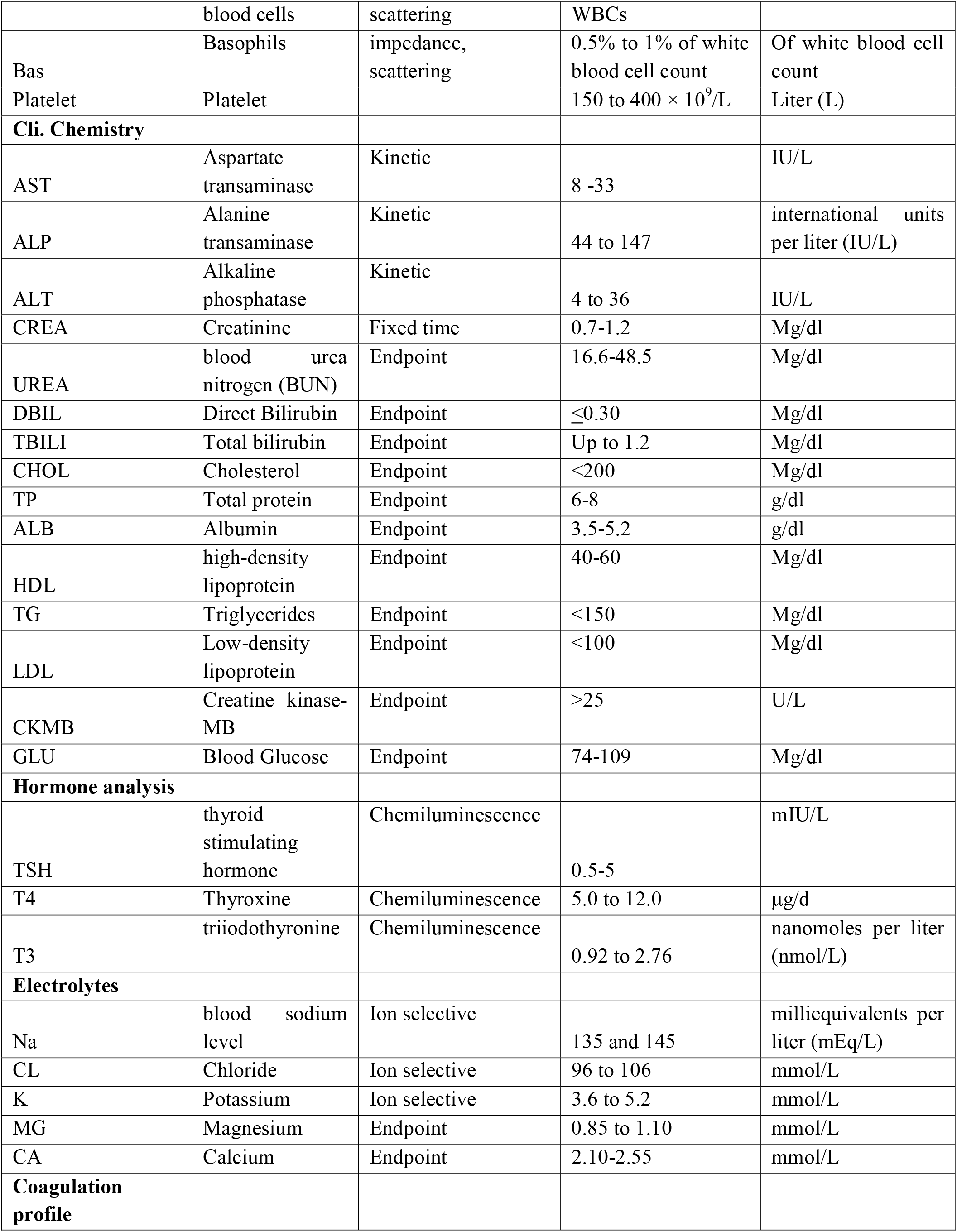
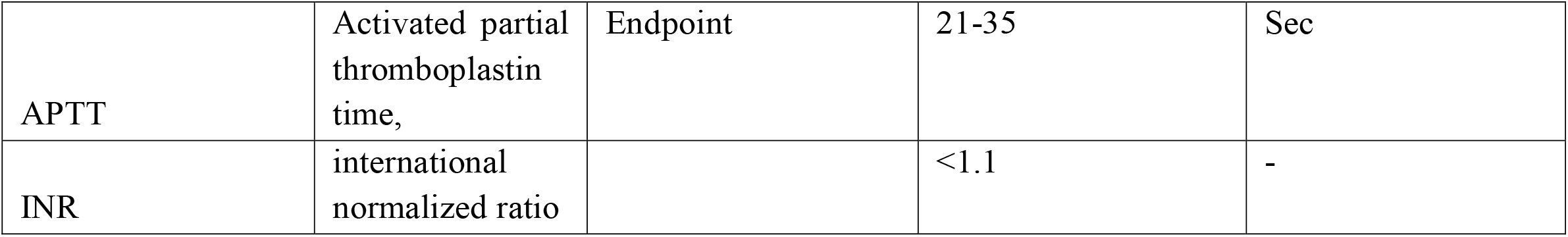
Laboratory parameters with the testing method used in this research.

**S3.** ROC analysis of promising biomarkers for severity of COVID-19.

**S3: ROC analysis of promising biomarkers for severity of COVID-19.**
| Predictor | direction | Optimum cutpoint | sensitivity | specificity | AUC |
| --- | --- | --- | --- | --- | --- |
| Wbc |  | 8.75 | 0.70 | 0.48 | 0.60 |
| Urea |  | 47.35 | 0.36 | 0.78 | 0.57 |

## Notes

### Competing Interest Statement

The authors have declared no competing interest.

### Author Declarations

The research protocol had been getting approval from different institutional ethics and research committees prior to the analysis. Accordingly it was approved by the IRB of the College of Health Sciences, Addis Ababa University IRB committee (IRB # 029/20/Lab) IRB of the Department of Medical Laboratory Sciences Addis Ababa University (reference #-MLS/174/21) IRB office of Addis Ababa City Administration Health Bureau, AAPHREML (Reference #- AAHB/4039/227) and also by Yekatit 12 Hospital Medical College (IRB protocol # 75/20) and also reviewed and approved by the Ekka Kotebe hospital IRB protocol # Eka-150-5-4). Study participants consented/assented in writing to sample storage and subsequent use of their samples and a waiver of consent was provided by the institutional review board for the use of leftover samples. Moreover this study was conducted per the Helsinki Declarations general guiding principles and regulations.

## Reference

1. WHO Coronavirus (COVID-19) Dashboard|WHO Coronavirus (COVID-19) Dashboard with Vaccination Data. Available online: https://covid19.who.int/. Accessed Oct 12, 2022.

2. University JH. Johns Hopkins University, J.H., worldometers COVID 19 Daily update. worldometers. Available from: https://coronavirus.jhu.edu/. Accessed Oct 12, 2022.

3. Abagero A, Ragazzoni L, Hubloue I, Barone-Adesi F, Lamine H, Addissie A, et al., A Review of COVID-19 Response Challenges in Ethiopia. International Journal of Environmental Research and Public Health. 2022; 19(17):11070. https://doi.org/10.3390/ijerph191711070

4. Niknam, Z., Jafari, A., Golchin, A. et al. Potential therapeutic options for COVID-19: an update on current evidence. Eur J Med Res 27, 6 (2022). https://doi.org/10.1186/s40001-021-00626-3

5. Battaglini D, Lopes-Pacheco M, Castro-Faria-Neto HC, Pelosi P and Rocco PRM (2022) Laboratory Biomarkers for Diagnosis and Prognosis in COVID-19. Front. Immunol. 13:857573. doi: 10.3389/fimmu.2022.857573

6. Leulseged TW, Hassen IS, Ayele BT, Tsegay YG, Abebe DS, Edo MG, et al. (2021) Laboratory biomarkers of COVID-19 disease severity and outcome: Findings from a developing country. PLoS ONE 16(3): e0246087. https://doi.org/10.1371/journal.pone.0246087

7. Bivona G, Agnello L, Ciaccio M. Biomarkers for Prognosis and Treatment Response in COVID-19 Patients. Ann Lab Med. 2021;41(6):540–548. doi:10.3343/alm.2021.41.6.540.

8. Samprathi M and Jayashree M (2021) Biomarkers in COVID-19: An Up-To-Date Review. Front. Pediatr. 8:607647. doi: 10.3389/fped.2020.607647

9. World Health Organization. Living guidance for clinical management of COVID-19. Geneva: 23 NOVEMBER 2021

10. Yang, Xiaobo, et al. “Clinical course and outcomes of critically ill patients with SARS-CoV-2 pneumonia in Wuhan, China: a single-centered, retrospective, observational study.” The Lancet Respiratory Medicine 8.5 (2020): 475–481,

11. Arentz M, Yim E, Klaff L, et al. Characteristics and Outcomes of 21 Critically Ill Patients With COVID-19 in Washington State. JAMA. 2020; 323(16):1612–1614. doi:10.1001/jama.2020.4326

12. Dongelmans, D.A., Termorshuizen, F., Brinkman, S. et al. Characteristics and outcome of COVID-19 patients admitted to the ICU: a nationwide cohort study on the comparison between the first and the consecutive upsurges of the second wave of the COVID-19 pandemic in the Netherlands. Ann. Intensive Care 12, 5 (2022). https://doi.org/10.1186/s13613-021-00978-3

13. Chen, Ruchong, et al. “Risk factors of fatal outcome in hospitalized subjects with coronavirus disease 2019 from a nationwide analysis in China.” Chest 158.1 (2020): 97–105

14. Khan, Anas, et al. “Risk factors associated with worse outcomes in COVID-19: a retrospective study in Saudi Arabia.” Eastern Mediterranean Health Journal 26.11 (2020).

15. Leulseged, Tigist W., et al. “Factors associated with development of symptomatic disease in Ethiopian COVID-19 patients: a case-control study.” BMC infectious diseases 21.1 (2021): 1–7.

16. Liang X. Is COVID-19 more severe in older men? Postgraduate Medical Journal 2020; 96:426.

17. Nizami DJ, Raman V, Paulose L, Hazari KS, Mallick AK. Role of laboratory biomarkers in assessing the severity of COVID-19 disease. A cross-sectional study. J Family Med Prim Care. 2021 Jun; 10(6):2209–2215. doi: 10.4103/jfmpc.jfmpc_145_21. Epub 2021 Jul 2. PMID: 34322414; PMCID: PMC8284239.

18. Yang, Hai-Jun, et al. “Predictors of mortality for patients with COVID-19 pneumonia caused by SARS-CoV-2.” European Respiratory Journal 56.3 (2020).]

19. De Meester, Johan, et al. “Incidence, characteristics, and outcome of COVID-19 in adults on kidney replacement therapy: a regionwide registry study.” Journal of the American Society of Nephrology 32.2 (2021): 385–396.

20. Zhou, Fei, et al. “Clinical course and risk factors for mortality of adult inpatients with COVID-19 in Wuhan, China: a retrospective cohort study.” The Lancet 395.10229 (2020): 1054–1062

